# Integration of MRI radiomics and germline genetics to predict the IDH mutation status of gliomas

**DOI:** 10.1101/2024.07.16.24310519

**Authors:** Taishi Nakase, George Henderson, Thomas Barba, Rohan Bareja, Geno Guerra, Qingyu Zhao, Stephen S. Francis, Olivier Gevaert, Linda Kachuri

**Affiliations:** Department of Epidemiology and Population Health, Stanford University School of Medicine, Stanford, CA, USA; Stanford Center for Biomedical Informatics Research (BMIR), Stanford University, Stanford, CA, USA; Department of Internal Medicine, Edouard Herriot Hospital, Lyon, France; Department of Neurological Surgery, University of California San Francisco, San Francisco, CA, USA; Department of Radiology, Weill Cornell Medicine, New York, NY, USA; Department of Epidemiology and Biostatistics, University of California San Francisco, San Francisco, CA, USA; Weill Institute for Neurosciences, University of California San Francisco, San Francisco, CA, USA; Department of Biomedical Data Science, Stanford University, Stanford, CA, USA; Stanford Cancer Institute, Stanford University School of Medicine, Stanford, CA, USA

## Abstract

The molecular profiling of gliomas for isocitrate dehydrogenase (*IDH*) mutations currently relies on resected tumor samples, highlighting the need for non-invasive, preoperative biomarkers. We investigated the integration of glioma polygenic risk scores (PRS) and radiographic features for prediction of *IDH* mutation status. We used 256 radiomic features, a glioma PRS and demographic information in 158 glioma cases within elastic net and neural network models. The integration of glioma PRS with radiomics increased the area under the receiver operating characteristic curve (AUC) for distinguishing IDH-wildtype vs. IDH-mutant glioma from 0.83 to 0.88 (P_ΔAUC_=6.9×10^-5^) in the elastic net model and from 0.91 to 0.92 (P_ΔAUC_=0.32) in the neural network model. Incorporating age at diagnosis and sex further improved the classifiers (elastic net: AUC=0.93, neural network: AUC=0.93). Patients predicted to have IDH-mutant vs. IDH-wildtype tumors had significantly lower mortality risk (hazard ratio (HR)=0.18, 95% CI: 0.08-0.40, P=2.1×10^-5^), comparable to prognostic trajectories for biopsy-confirmed *IDH* status. The augmentation of imaging-based classifiers with genetic risk profiles may help delineate molecular subtypes and improve the timely, non-invasive clinical assessment of glioma patients.

## INTRODUCTION

Gliomas are the most common primary malignant brain tumors in adults^1^. These neoplasms encompass multiple subtypes with distinct somatic mutations that delineate different clinical trajectories^2^. Although glioma classifications continue to evolve, several key features have been used to define molecular subtypes since 2016: isocitrate dehydrogenase 1 and 2 mutations (collectively referred to as *IDH* mutations), chromosome 1p and 19q co-deletion, and *TERT* promoter mutations^2–4^. The 2021 World Health Organization (WHO) glioma classification guidelines use these tumor molecular features to define three glioma subtypes^5^: tumors without *IDH* mutation (IDH-wildtype glioblastomas), tumors with *IDH* mutation and an unbalanced translocation between chromosomes 1 and 19 (IDH-mutant 1p19q co-deleted oligodendrogliomas), and IDH-mutant tumors without 1p19q co-deletion (IDH-mutant astrocytomas). IDH-wildtype glioblastomas (GBM) are more aggressive, have fewer treatment options and are associated with significantly shorter overall survival than IDH-mutant gliomas^2,6^. The early establishment of a molecular diagnosis for gliomas is important to predict tumor behavior and guide treatment of individual patients^7–9^.

Currently, the classification of gliomas into prognostically significant subtypes is based on histopathological and molecular assessment of tissue samples obtained from biopsy or resection. Therefore, the evaluation of treatment options and prognostication are often delayed until after surgery, which carries the risk of permanent operative complications or may not be readily accessible in low-resource settings. Recent efforts have been aimed at using noninvasive procedures such as imaging^10–13^ and germline genotyping^14,15^ to provide insight into the presence or absence of clinically relevant somatic mutations (e.g. *IDH* mutation) prior to surgical interventions. By providing timely insight into the tumor molecular profile, these noninvasive tools may complement the standard histopathological assessment of surgical specimens to expedite treatment decisions and better inform patient management.

Major advances have been made in the use of tumor radiographic features obtained from preoperative imaging data to classify gliomas into molecular subtypes. Earlier applications of machine learning (ML) to imaging data for tumor classification used textural analysis approaches or rule-based systems such as VASARI^16,17^. Since these approaches rely on manual feature selection, research efforts have since focused on the development of deep learning models such as convolutional neural networks (CNNs) that automatically extract features from complex images^11–13^. Recent CNN-based models for glioma classification have shown promising results, with model predictions recapitulating prognostic outcomes expected for different molecular subtypes. However, previous studies of imaging features have not accounted for other known clinical and genetic indicators of subtype-specific glioma risk.

Genome-wide association studies (GWAS) for glioma have shown that inherited genetic variation influences disease risk and that different molecular subtypes are associated with distinct genetic risk loci^18,19^. Polygenic risk scores (PRS), which aggregate the effects of risk alleles across the genome to provide a personalized genetic susceptibility profile^20^, have been shown to predict subtype-specific glioma risk and accurately distinguish among molecular subtypes^14,15^. PRS require genotyping or sequencing of constitutive DNA obtained from buccal cells or blood samples and can thus provide a non-invasive ancillary measure of subtype-specific glioma risk prior to surgery. Since gliomas sometimes present with non-characteristic radiographic features (e.g. IDH-wildtype non-enhancing tumors)^13^, inherited genetic variation could offer an additional indicator of tumor aggressiveness (e.g. *IDH* mutation status) independent of radiomic features that might improve the performance of imaging-based classification models.

In this study, we integrated radiomic features extracted from preoperative multimodal magnetic resonance imaging (MRI) scans with germline PRS profiles to classify gliomas according to *IDH* mutation status. Using the developed classification model, we also identify predictive features in subtype discrimination and assess their clinical significance.

## METHODS

### Study population

The analysis group consisted of 768 primary glioma cases (384 IDH-wildtype, 384 IDH-mutant) with available tumor molecular data from The Cancer Genome Atlas (TCGA). TCGA is a public data repository for de-identified tumor samples from consented patients (https://www.cancer.gov/tcga). Analyses were restricted to individuals of predominantly European ancestry. Of the 768 glioma cases, 610 had genotyping data only and 158 (81 IDH-wildtype, 77 IDH-mutant) had both genotyping data and preoperative MRI scans available on the Cancer Imaging Archive (https://www.cancerimagingarchive.net)^21^. An overview of the study design and analysis is provided in **Figure 1**.

**Figure 1:**
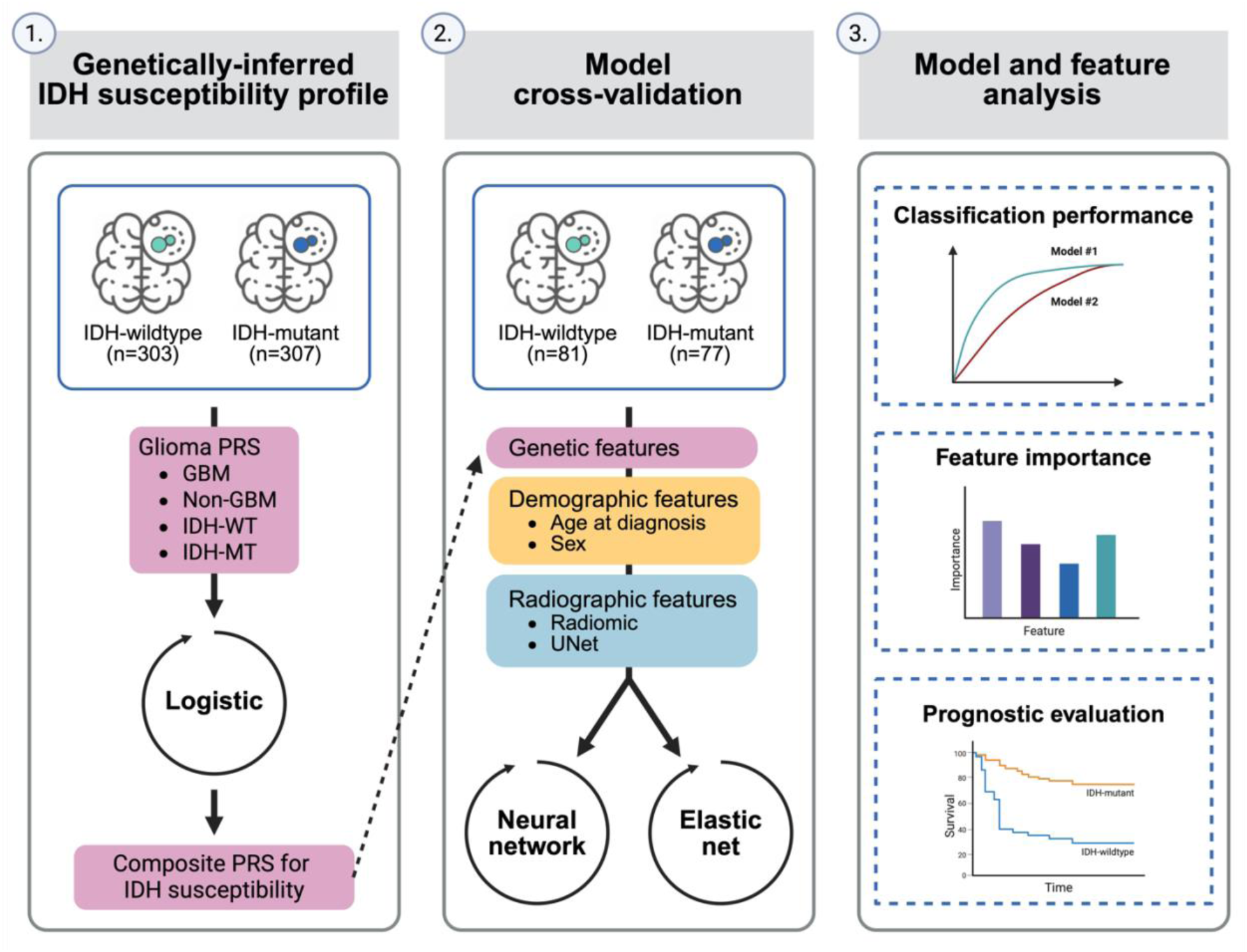
Overview of the study design. Abbreviations: PRS=polygenic risk score, GBM=glioblastoma, IDH-WT=IDH-wildtype, IDH-MT=IDH-mutant.

### Germline genetic predictors

All 768 cases were genotyped on the Affymetrix 6.0 array and imputed with the TOPMed reference panel, with standard quality control procedures as previously described^14,22^. Briefly, SNPs with a call rate <95% were excluded along with those at a low minor allele frequency (MAF<0.005) or showing significant deviation from the Hardy-Weinberg equilibrium (P<10^-6^). Using individual-level genotyping data, we fit four previously developed subtype-specific PRS for each patient in TCGA, as described in Nakase et al^14^. Briefly, the GBM PRS and the non-GBM PRS were trained using summary statistics from a GWAS of 10,346 cases (5395 GBM, 4466 non-GBM) and 14,687 controls, while PRS for molecular subtypes were developed using GWAS results from 2632 cases (1115 IDH-wildtype, 699 IDH-mutant) and 2445 controls^14,18,19^.

For each individual, each subtype-specific PRS was converted to a standardized z-score based on the in-sample TCGA distribution. To adjust for population stratification, we regressed out the effects of the first 10 genetic ancestry principal components and used the residualized z-scores for each PRS in subsequent processing. Next, we fit a logistic regression model with *IDH* mutation status as the outcome and the four residualized PRS features as the explanatory variables on the subset of 609 patients without radiomic data. This model was then applied to the 158 patients with both radiomic and germline genetic data to extract a new composite PRS feature based on the weighted effect of each subtype-specific PRS.

### Radiographic predictors

#### Radiomic features

Radiomic features extracted from the 158 cases with available preoperative MRI scans (T1-weighted pre-contrast [T1], T1-weighted post-contrast [T1-Gd], T2 and T2-FLAIR) were obtained from Bakas et al^23–25^. As described elsewhere^23^, MRI scans of each patient underwent standard pre-processing including registration, resampling and skull stripping, followed by computer-aided assignment of segmentation labels to tumor sub-regions (e.g. peritumoral edema). Computer-aided segmentation labels were then manually-revised by a neuroradiologist. Based on the assigned labels of each tumor sub-region, a panel of radiomic features were extracted, which included intensity, volumetric, morphologic, histogram, textural, spatial and tumor diffusion parameters. Of the 723 radiomic features, 467 were not available for all cases and were excluded from the analysis. For each of the remaining 256 radiomic features, we calculated its standardized z-score. We assessed for potential confounding by age, sex and brain volume (excluding skeletal structures) by calculating the Pearson correlation between the standardized z-score of each radiomic feature and each factor. For each feature that was significantly correlated with age, sex or brain volume (P<0.05), residualization was performed to omit the effect of these factors^26^.

#### UNet-based autoencoder features

UNet-based autoencoder (UNET) features extracted from the 158 cases with available preoperative MRI scans (T1, T1-Gd, FLAIR) were obtained from Barba et al^27^. Briefly, a 3D UNet model without skip connections was trained to encode and reconstruct sequence-specific brain MRI scans using images from 1600 healthy brains^28^ and 2214 morphologically abnormal brains with glioma^29,30^. The trained UNet-based autoencoder was then applied to the preprocessed MRI scans (e.g. registration, normalization, skull stripping) of the 158 glioma cases in TCIA to extract low-dimensional embeddings from the autoencoder’s bottleneck layer. The sequence-specific features were then concatenated and normalized yielding 6144 embeddings for each case. To reduce dimensionality while preserving quality, principal component analysis was applied to extract the first 96 principal components (PCs) that account for >90% of the explained variance.

### Model development and evaluation

We used an elastic net or a neural network to classify primary glioma cases according to *IDH* mutation status in the subset of the TCGA dataset with both radiomic and germline genetic information (n=158). Model features included age as diagnosis, sex, a composite PRS and either radiomic features or UNET features. The fully connected neural networks had two linear layers (size: 8, 16, 32, or 64) and dropout following each linear layer (probability: 0, 0.1, or 0.2), with selection of model parameters based on binary cross entropy loss across 240 training epochs. Standard logistic regression was used for the demographics-only (age at diagnosis and sex) and PRS-only models.

To reduce bias in estimates of model performance, we performed nested cross-validation (CV), as described in **Supplementary Figure 1**. Briefly, the 158 cases from the TCGA dataset were partitioned into 5 outer CV folds with each fold divided into a training set (80%) and a hold-out test set (20%). For each outer training fold, we performed 5-fold CV to tune the hyperparameters and then fitted the tuned model to the whole training fold. The fitted model with the best hyperparameter configuration was then applied to the corresponding hold-out test set. Model performance was evaluated across the predictions of the concatenated hold-out test sets of each of the 5 outer CV folds. Classification performance was quantified using accuracy, precision, recall, and area under the receiver operating characteristic curve (AUC). The nested cross-validation procedure was repeated for 500 random training/testing splits to obtain the distribution of each of the performance metrics. Feature importance was quantified using the distribution of weights in the elastic net model with demographic, PRS and radiomics features across the 500 random iterations of the nested CV procedure.

### Survival analysis

We examined how well the demographic, genetic and radiographic features predicted overall survival by IDH status, using the full integrated elastic net classifier. Follow-up time was calculated from date of diagnosis to death or end of follow-up. Differences in survival trajectories were visualized using Kaplan-Meier and tested using the log rank test. To incorporate covariates, hazard ratios (HR) for predicted IDH status were estimated using Cox proportional hazards models with adjustment for age at diagnosis, sex and the first 10 genetic ancestry principal components. We also evaluated the association of each feature that was included in the classifier with overall survival.

## RESULTS

Basic summary information of study participants overall and stratified by available genotyping and radiomic data is provided in **Table 1**. The median age of included cases was 52 years, with more males (58.5%) than females (41.5%). Overall, we included 384 subjects diagnosed with IDH-wildtype glioma and 384 with IDH-mutant glioma.

**Table 1:**
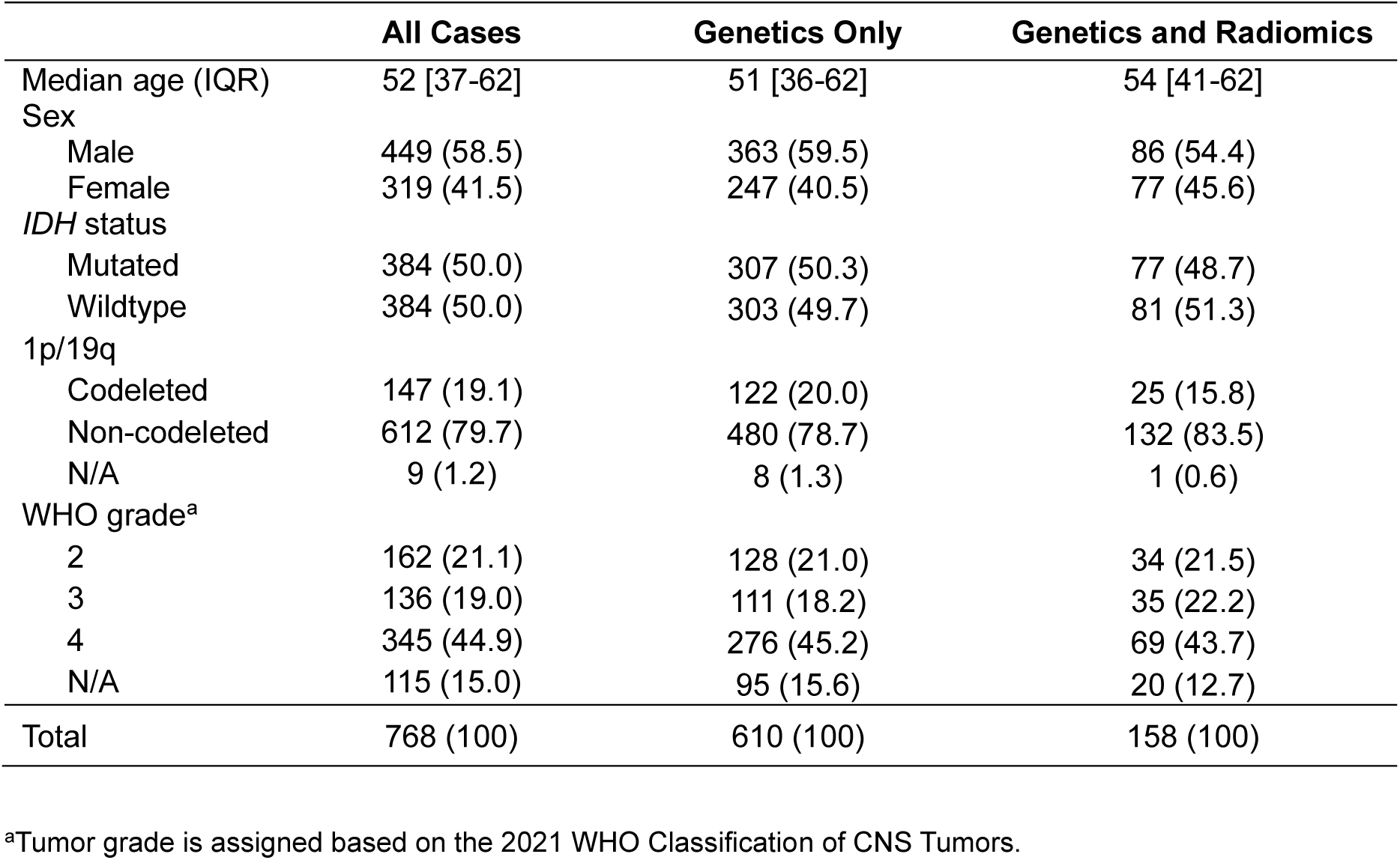
Characteristics of glioma cases in The Cancer Genome Atlas (TCGA). Characteristics of glioma cases in TCGA are stratified by available germline genetic and radiomic data. For categorical variables, percentage (%) is reported. For quantitative variables, interquartile range (IQR) is reported.

### Evaluation of multi-modal *IDH* classifiers

In elastic net models limited to features of a single class (i.e. demographic [age and sex], genetic or radiographic), we found that the radiomics-based model was the most predictive for *IDH* status classification (**Table 2, Supplementary Figure 2**). Radiomic features yielded the highest mean AUC (0.83, 95% confidence interval [CI]: 0.79-0.86), followed by the demographics-only model (0.76, 95% CI: 0.74-0.77), the UNET-PC model (0.72, 95% CI: 0.64-0.78) and the composite PRS model (0.68, 95% CI: 0.66-0.70). Radiomic features also exhibited improved accuracy (0.80), precision (0.82) and recall (0.77) compared to the other single-class models.

**Table 2:**
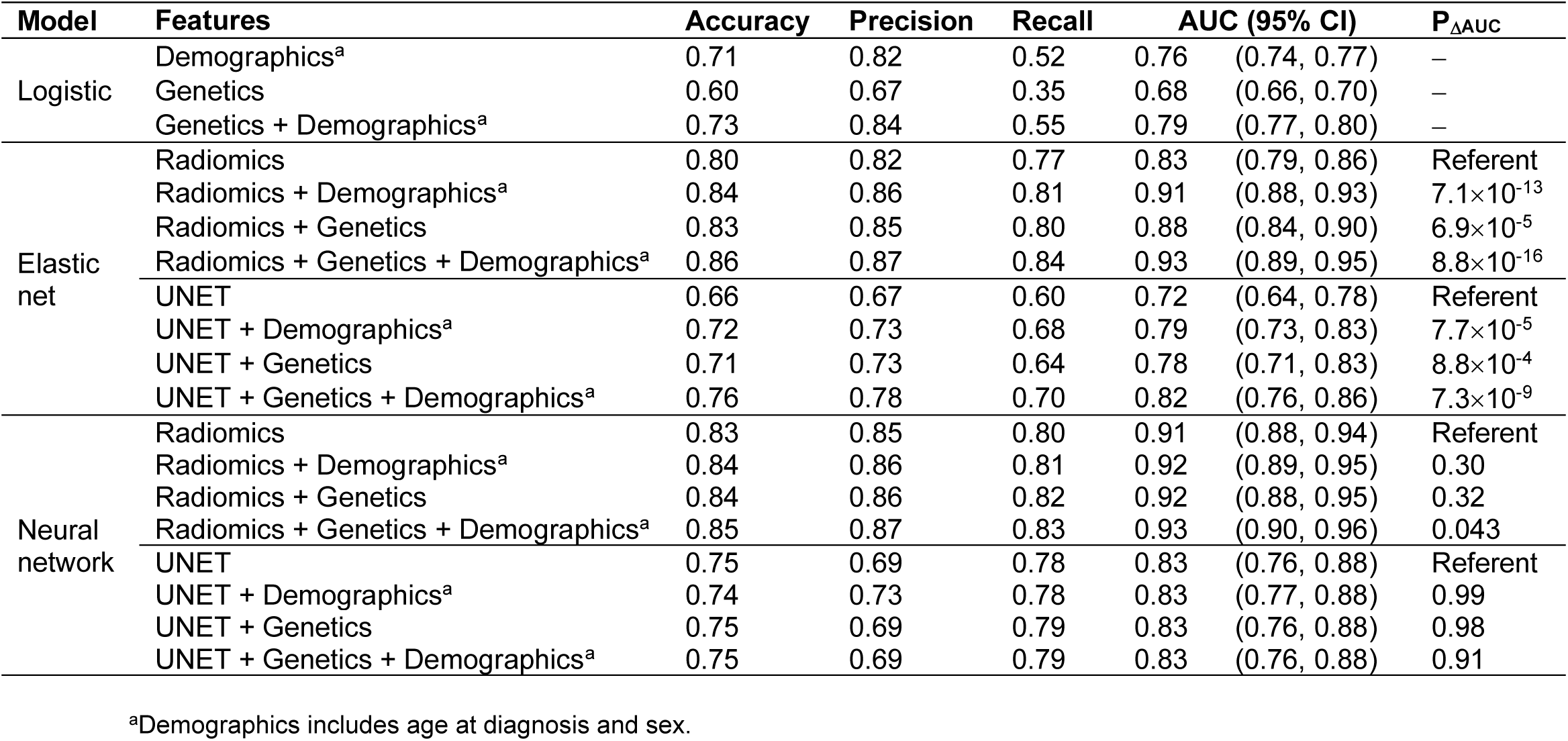
Classification performance for IDH mutation status. Performance metrics are based on classification models for *IDH* mutation status in glioma cases. Models with radiomic features and UNet-based autoencoder (UNET) features were trained with an elastic net or neural network; otherwise, models were trained with logistic regression. The mean estimate for accuracy, precision, recall and AUC across 500 random iterations of the nested cross-validation procedure are provided. Two-sided p-values (P_ΔAUC_) testing the differences in AUC between the radiographic-only model (radiomics [elastic net], UNET [elastic net], radiomics [neural network] and UNET [neural network]) and the integrated models are based on a resampled t-test^35,36^.

| Model | Features | Accuracy | Precision | Recall | AUC (95% CI) | $P_{\Delta AUC}$ |
| --- | --- | --- | --- | --- | --- | --- |
| Logistic | Demographics<sup>a</sup> | 0.71 | 0.82 | 0.52 | 0.76 (0.74, 0.77) | – |
|  | Genetics | 0.60 | 0.67 | 0.35 | 0.68 (0.66, 0.70) | – |
|  | Genetics + Demographics<sup>a</sup> | 0.73 | 0.84 | 0.55 | 0.79 (0.77, 0.80) | – |
| Elastic net | Radiomics | 0.80 | 0.82 | 0.77 | 0.83 (0.79, 0.86) | Referent |
| | Radiomics + Demographics<sup>a</sup> | 0.84 | 0.86 | 0.81 | 0.91 (0.88, 0.93) | $7.1 \times 10^{-13}$ |
| | Radiomics + Genetics | 0.83 | 0.85 | 0.80 | 0.88 (0.84, 0.90) | $6.9 \times 10^{-5}$ |
| | Radiomics + Genetics + Demographics<sup>a</sup> | 0.86 | 0.87 | 0.84 | 0.93 (0.89, 0.95) | $8.8 \times 10^{-16}$ |
|  | UNET | 0.66 | 0.67 | 0.60 | 0.72 (0.64, 0.78) | Referent |
| | UNET + Demographics<sup>a</sup> | 0.72 | 0.73 | 0.68 | 0.79 (0.73, 0.83) | $7.7 \times 10^{-5}$ |
| | UNET + Genetics | 0.71 | 0.73 | 0.64 | 0.78 (0.71, 0.83) | $8.8 \times 10^{-4}$ |
| | UNET + Genetics + Demographics<sup>a</sup> | 0.76 | 0.78 | 0.70 | 0.82 (0.76, 0.86) | $7.3 \times 10^{-9}$ |
| Neural network | Radiomics | 0.83 | 0.85 | 0.80 | 0.91 (0.88, 0.94) | Referent |
|  | Radiomics + Demographics<sup>a</sup> | 0.84 | 0.86 | 0.81 | 0.92 (0.89, 0.95) | 0.30 |
|  | Radiomics + Genetics | 0.84 | 0.86 | 0.82 | 0.92 (0.88, 0.95) | 0.32 |
|  | Radiomics + Genetics + Demographics<sup>a</sup> | 0.85 | 0.87 | 0.83 | 0.93 (0.90, 0.96) | 0.043 |
|  | UNET | 0.75 | 0.69 | 0.78 | 0.83 (0.76, 0.88) | Referent |
|  | UNET + Demographics<sup>a</sup> | 0.74 | 0.73 | 0.78 | 0.83 (0.77, 0.88) | 0.99 |
|  | UNET + Genetics | 0.75 | 0.69 | 0.79 | 0.83 (0.76, 0.88) | 0.98 |
|  | UNET + Genetics + Demographics<sup>a</sup> | 0.75 | 0.69 | 0.79 | 0.83 (0.76, 0.88) | 0.91 |

<sup>a</sup>Demographics includes age at diagnosis and sex.

Combining germline genetic, demographic, and radiographic features improved classification of *IDH* status compared to each class of predictors alone (**Table 2**, **Supplementary Figure 3**). PRS and radiomic features had significantly improved performance compared to radiomics alone, with a mean AUC of 0.88 (95% CI: 0.84-0.90, P_ΔAUC_=6.9×10^-5^). Classification of *IDH* status was further improved by adding age at diagnosis and sex (AUC=0.91, 95% CI: 0.88-0.93). Overall, the model that included age, sex, radiomic features, and PRS achieved the highest mean AUC (0.93, 95% CI: 0.89-0.95), accuracy (0.87), precision (0.84) and recall (0.84) (**Figure 2A**). Although performance was attenuated using UNET-PC features compared to radiomic features, we found that incorporating PRS and demographic information increased the mean AUC from 0.72 (95% CI: 0.64-0.78) to 0.82 (95% CI: 0.76-0.86, P_ΔAUC_=7.3×10^-9^) (**Table 2**, **Figure 2B**).

**Figure 2:**
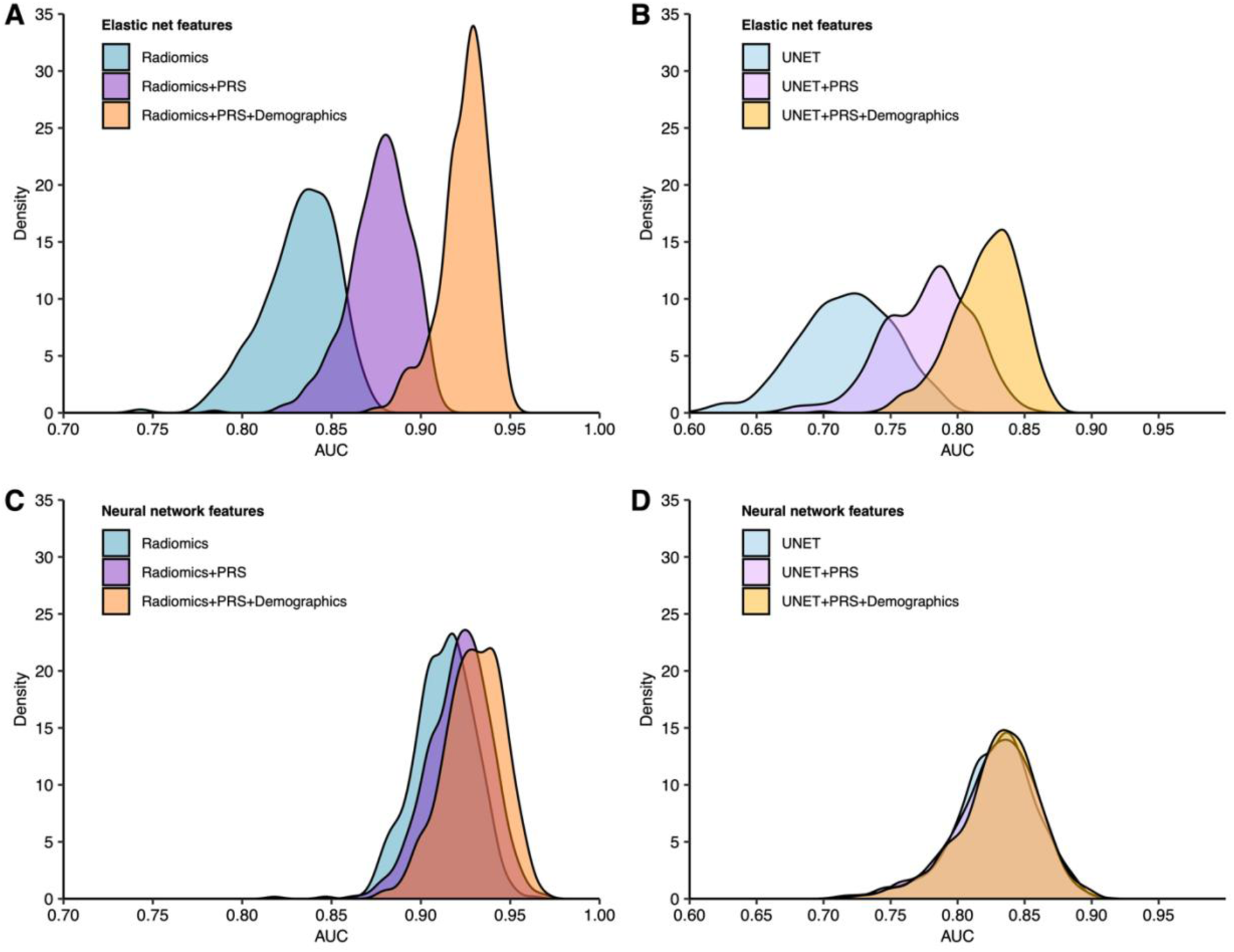
Distribution of AUC for classification of IDH mutation status from repeated nested cross-validation. AUC estimates from 500 random iterations of nested cross-validation were obtained from IDH classification models using different combinations of features (radiomics/UNET, PRS, demographics [age, sex]) and model architectures (elastic net, neural network).

In sensitivity analyses using radiomic features that were not adjusted for confounding factors such as age, sex and brain volume, we found that multimodal classifiers that integrated demographic information and PRS with radiomic features showed the best performance (**Supplementary Figure 4**, **Supplementary Table 1**). Compared to the radiomics-only model (AUC=0.89, 95% CI: 0.88-0.90), the combined radiomics and PRS model (AUC=0.92, 95% CI: 0.89-0.94, P_ΔAUC_=6.6×10^-5^) and the full integrated model (AUC=0.93, 95% 95% CI: 0.90-0.94, P_ΔAUC_=1.2×10^-6^) showed significantly improved performance.

Next, we evaluated multimodal classifiers for *IDH* status trained using a neural network architecture that accommodates more complex, non-linear interactions between predictors (**Table 2**). Combining PRS with radiomic features did not produce an appreciable increase in mean AUC (0.92, 95% CI: 0.88-0.95) relative to the radiomics-only model (AUC=0.91, 95% CI: 0.88-0.94). However, the integration of all three data modalities demonstrated a modestly higher mean AUC of 0.93 (95% CI: 0.90-0.96, P_ΔAUC_=0.043) compared to radiomics alone (**Figure 2C**). The integration of genetic and demographic information with UNET features in a neural network model did not alter classification performance (AUC=0.83, **Figure 2D**).

### Feature importance

Next, we sought to gain more insight into factors contributing to model performance by examining feature weights across all predictors in the elastic net model. Overall, 40 out of the 256 features had non-zero cumulative weights in >50% of the 500 random iterations of nested cross-validation (**Figure 3A**). Of those 40 features, 11 were predictive of *IDH* mutation status in all 500 iterations: age at diagnosis, the composite PRS, the ratio of the enhancing tumor (ET) volume to whole tumor (WT) volume, the percentage of the tumor core (TC) in the frontal lobe, the ratio of the non-enhancing tumor (NET) volume to the WT volume, the ratio of the ET to the TC volume, the ratio of the NET volume to the TC volume and several metrics of the heterogeneity of the NET (**Figure 3B**). Compared to IDH-wildtype tumors, IDH-mutant tumors were generally diagnosed at an earlier age, had minimal areas of enhancement and more often developed in the frontal lobe (**Figure 3B**).

**Figure 3:**
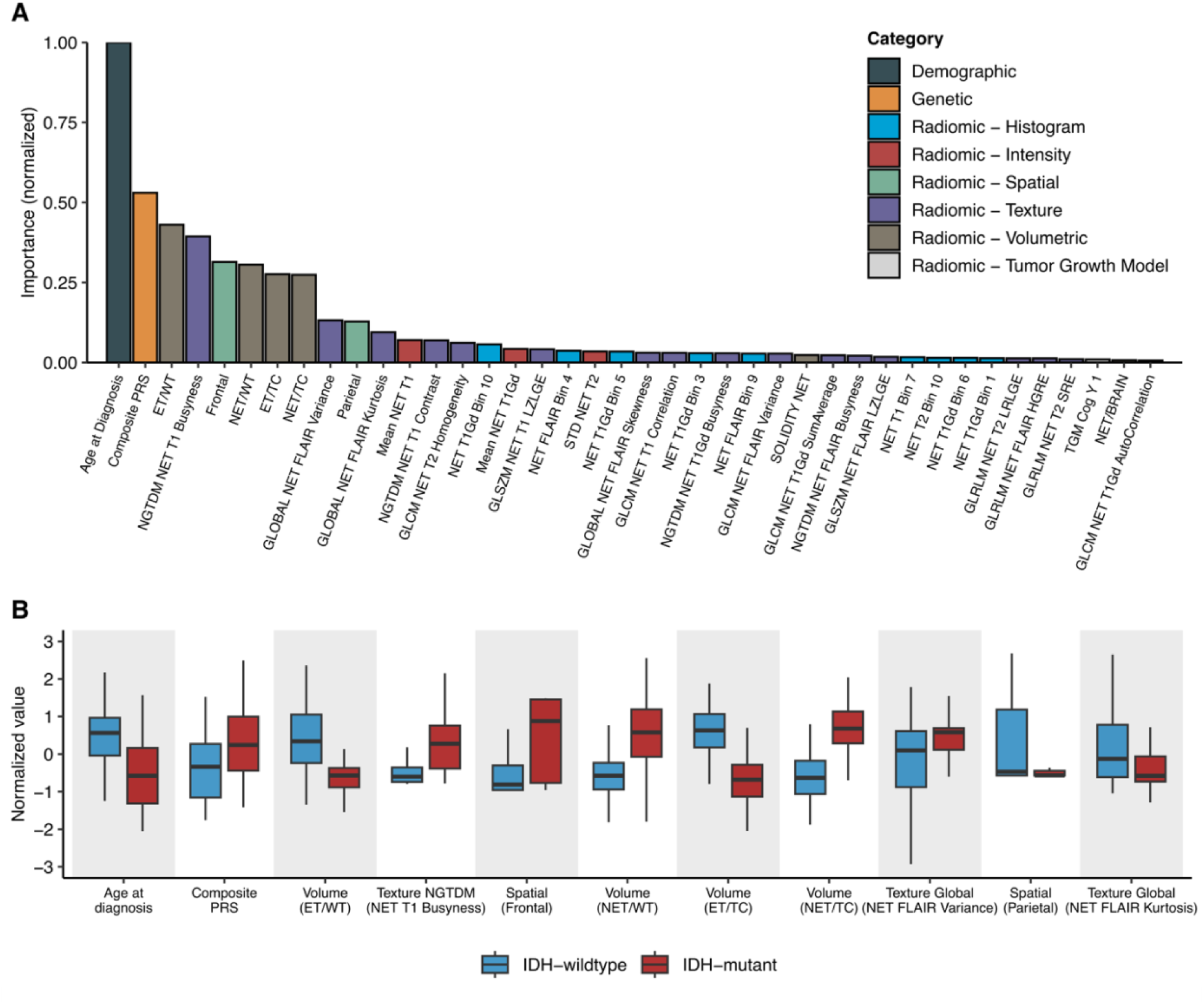
Predictive features for IDH status classification. (**A)** Mean cumulative value of feature weight in the full integrated elastic net model across the 500 random iterations of nested cross-validation. Feature weights are normalized by the maximum value. Features with non-zero cumulative weights in >50% of the 500 random iterations are shown. (**B**) Distribution of the predictive features stratified by tumor subtype. Features were defined as predictive if the cumulative weight was non-zero across all 500 random iterations of nested cross-validation. Features are ordered (from left to right) in decreasing importance. Abbreviations: NET=Non-enhancing tumor, WT=Whole tumor, TC=Tumor core, ET=Enhancing tumor, GLCM= Grey-level co-occurrence matrix, GLSZM=Gray-level size zone matrix, GLRLM= Gray-level run-length matrix, SZLGE=Small zone low gray-level emphasis, SRE=Short run emphasis, NGTDM=Neighborhood grey-tone difference matrix.

### Multimodal *IDH* classifier predicts mortality

To evaluate the potential utility of our multimodal *IDH* classifier, we compared its association with mortality to ground-truth labels based on confirmed IDH status. Median survival time for patients predicted to have IDH-mutant (87.4 months) vs. IDH-wildtype tumors (14.7 months) was similar in magnitude to the survival differences observed based on biopsy-confirmed IDH mutation status (87.4 months vs. 14.1 months; **Figure 4A**). The difference in median survival between predicted IDH-mutant and predicted IDH-wildtype was 72.7 months (P=6.8×10^-11^) (**Figure 4B**). Associations with mortality for predicted IDH-mutant status remained robust in multivariable Cox models adjusted for genetic ancestry PCs and demographic information (HR=0.18, 95% CI: 0.08-0.40, P=2.1×10^-5^) (**Table 3**).

**Figure 4:**
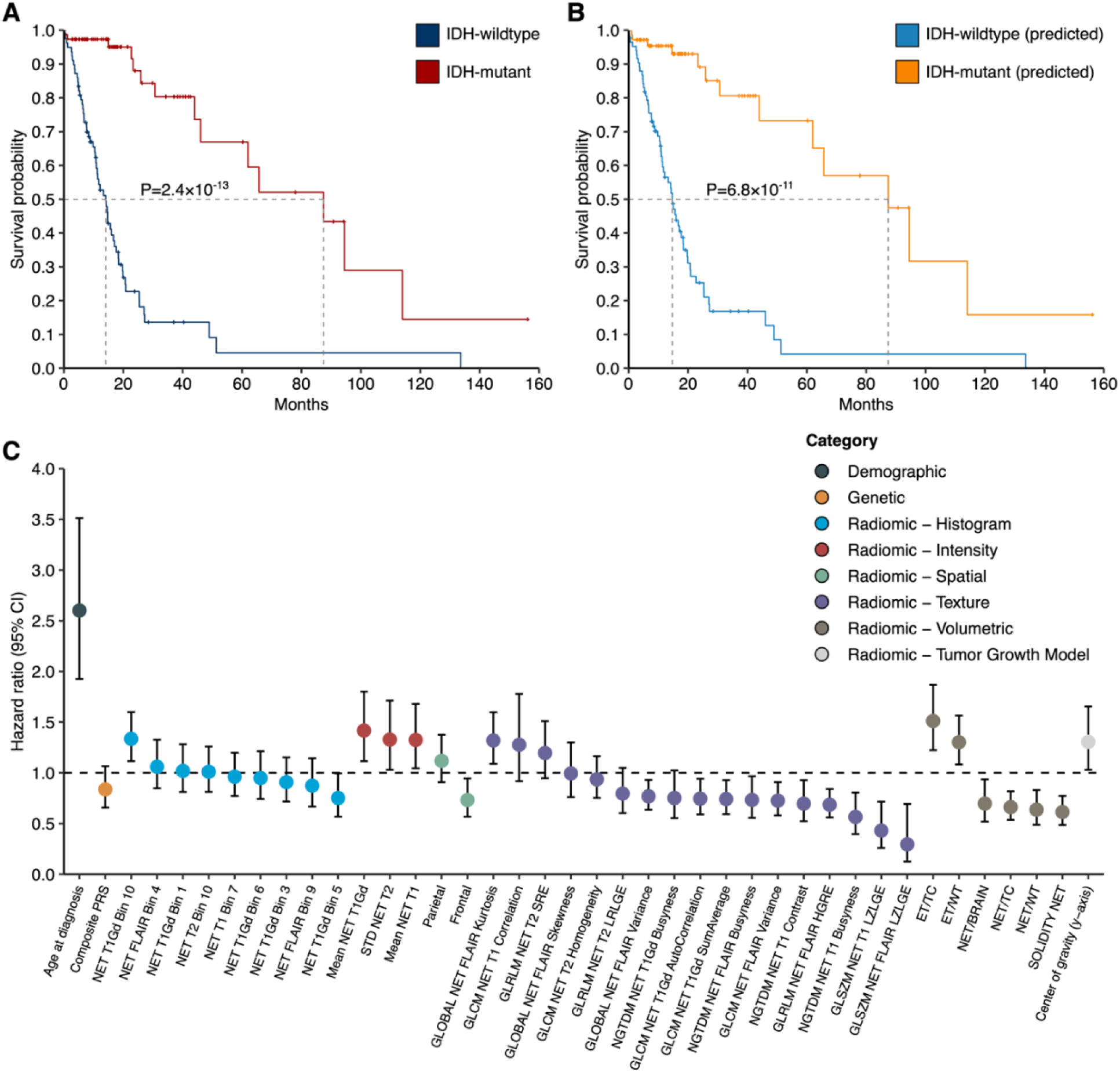
Overall survival for adult glioma cases with predicted or biopsy-confirmed IDH mutation status. (**A, B**) Percent survival distributions for adults with glioma stratified by predicted or biopsy-confirmed IDH mutation status. Differences in event time distributions were assessed using the log rank test. (**A**) IDH mutation status confirmed by molecular profiling of tumor samples. (**B**) IDH mutation status predicted based on integrated elastic net model (“predicted”). (**C**) Features used for IDH status classification are examined for association with overall survival using univariate Cox proportional hazards regression. Hazard ratio (HR) and 95% confidence interval (CI) for each feature are presented. Abbreviations: NET=Non-enhancing tumor, WT=Whole tumor, TC=Tumor core, ET=Enhancing tumor, GLCM= Grey-level co-occurrence matrix, GLSZM=Gray-level size zone matrix, GLRLM= Gray-level run-length matrix, SZLGE=Small zone low gray-level emphasis, SRE=Short run emphasis, NGTDM=Neighborhood grey-tone difference matrix

**Table 3:** Association between IDH mutation status and survival among glioma cases. Hazard ratios (HR) and 95% confidence intervals (CI) were estimated using Cox regression models adjusted for age at diagnosis, sex and genetic ancestry principal components. Predicted subtype labels are based on the full elastic net model with radiomic, germline genetic and demographic features, while ground truth subtype labels are based on molecular profiling of biopsied tumor samples.

|  | **Cases**<br>**(Deaths)** | **Median Survival**<br>**(Months)** | **HR (95% CI)** | **P value** |
| --- | --- | --- | --- | --- |
| Predictions |  |  |  |  |
| IDH-wildtype | 85 (57) | 14.7 | 1.00 |  |
| IDH-mutant | 73 (15) | 87.4 | 0.18 (0.08, 0.40) | 2.1×10<sup>-5</sup> |
| Ground truth |  |  |  |  |
| IDH-wildtype | 81 (58) | 14.1 | 1.00 |  |
| IDH-mutant | 77 (14) | 87.4 | 0.17 (0.08, 0.36) | 4.3×10<sup>-6</sup> |

When considering individual predictors, of the 40 features that had non-zero cumulative weights in >50% of the 500 random iterations of the integrated elastic net classifier, age at diagnosis and 24 radiomic features were associated with all-cause mortality at P<0.05 (**Figure 4C, Supplementary Table 2**). The composite PRS predictor was not strongly predictive of mortality (HR=0.84, 95% CI: 0.66-1.07). Prognostic radiomic features were primarily related to characteristics of the ET and NET regions. For instance, higher relative enhancing tumor volume (i.e. ET/TC) was associated with a 51% increase in overall mortality risk (HR=1.51, 95% CI: 1.22-1.87, P=1.3×10^-4^), while tumors with greater solidity of the NET had lower mortality risk (HR=0.61, 95% CI: 0.49-0.77, P=3.3×10^-5^).

## DISCUSSION

Molecular tumor markers, especially *IDH* mutation status, provide critical information for prognosis^7,8^ and treatment response^9^. Currently, the classification of gliomas into molecular subtypes relies on immunohistochemical profiling or sequencing of surgically resected tumor specimens, which often delays the evaluation of non-surgical treatment options. In this case study, we integrated radiographic features extracted from preoperative MRI scans with a composite polygenic risk score (PRS) for risk of IDH-mutant glioma, to examine the potential of germline genetic information to improve non-invasive, preoperative determination of *IDH* mutation status in glioma patients.

Previous models used to classify gliomas into clinically-relevant molecular subtypes have relied on either radiographic features^11–13^ or germline genetic markers^14,15^, but have not considered PRS or integrated germline genetic risk profiles with other modalities. Compared to single variants with small effects, PRS is a more powerful predictor that aggregates genetic susceptibility signals across the entire genome. The composite PRS evaluated in this study was trained using the largest available GWAS of glioma subtypes that did not include TCGA^18,19^. We found that the inclusion of diverse features extracted from multiple complementary modalities improved the ability to preoperatively distinguish *IDH*-mutant from *IDH*-wildtype tumors. The models that included all available features achieved the best overall classification performance, with a mean AUC of 0.93 for both the elastic net and the neural network on repeated cross-validation. These results are consistent with observations for other cancers such as thyroid cancer, where PRS improved imaging-based classifiers of malignancy risk^31^. We also found that age at diagnosis and the composite PRS were among the most predictive features along with volume of ET and NET, and the percentage of the TC in the frontal lobe. Additionally, our predicted subtype labels reproduced prognostic associations that were statistically significant and concordant in magnitude to ground-truth *IDH* mutation status.

Multi-modal classifiers have the potential to improve clinical management by prioritizing patients suitable for neoadjuvant therapy with *IDH* inhibitors and avoiding delays in the use of adjuvant chemoradiotherapy. While combined temozolomide and radiotherapy remains the standard of care for IDH-wildtype glioblastoma^32^, Vorasenib, an inhibitor of *IDH1* and *IDH2* enzymes, was recently shown to improve progression-free survival in patients with IDH-mutant glioma^33^. Although the optimal timing of Vorasenib treatment has not been studied, when provided earlier in the disease course, it might delay subsequent interventions in patients with low-grade glioma and improve quality of life. However, the added value of ML classifiers in patient management will require careful evaluation of their potential risks and contextualization in different clinical settings.

This work has several limitations. First, statistical power for model comparisons and survival analyses was limited since only 21% of TCGA participants had both radiomic and genotyping data available. Second, while nested cross-validation provides some robustness by evaluating performance based on in-sample hold-out subsets, it can overfit the training data and result in inflated estimates of model performance. This is an important consideration for the neural network models, which are particularly vulnerable to overfitting on small sample sizes due to their large parameter space. Given the absence of additional independent cohorts that include all required data modalities (germline genotyping, molecular tumor profiling, imaging), the absolute value of our performance metrics should be interpreted with caution. Our results serve as a proof-of-concept of the potential added value of germline genetic information for preoperative classification of gliomas. Future collaborative efforts to develop resources across different clinical centers are required to facilitate more rigorous and unbiased evaluation of multimodal classifiers. Moreover, the lower AUC of models based on UNet-based radiographic features relative to radiomic features may in part be attributable to an imbalance between the large UNET feature space (n=6144) and the small sample size (n=158). Indeed, dimensionality reduction of the UNET features with principal component analysis was required to improve performance, suggesting that sample size limitations may have constrained the performance of the UNET features. Lastly, our analyses focused on *IDH* mutation status, and we did not consider more refined molecular subtypes based on additional features, such as 1p19q codeletion, *TERT* promoter mutations and *EGFR* amplification due to limited sample size. However, of the currently used somatic mutations, *IDH* status is the most prognostically significant^2,4^.

Moreover, the best-performing radiomic features were based on a predefined set of tumor-related variables from imaging data with manually-revised segmentation labels^23^. Several recent studies have developed end-to-end CNN-based models that perform tumor segmentation, unsupervised feature learning and classification within a single framework^11–13^. These multi-task deep learning approaches do not rely on manual delineation of tumor regions, reduce computational burden and require minimal input from health care providers once trained, thereby facilitating their integration into existing neuro-oncology workflows. However, current end-to-end classification models are mostly image-intensity based and do not account for other types of features that might be informative for subtype discrimination, which may limit their performance, especially in patients with non-characteristic radiographic findings. As individual-level genotyping on larger cohorts of glioma patients become available, multi-task deep learning models that incorporate germline genetic and demographic information can be trained using a late-fusion strategy^13,34^.

This work has several important strengths. Our study applies the best performing PRS for glioma that has been shown to accurately estimate subtype-specific glioma risk and distinguish glioma subtypes in external validation analyses^14^. We leveraged genetic data without corresponding radiomic data in TCGA to generate a composite PRS feature that reflects the joint effects of multiple subtype-specific PRS. In addition to evaluating classification of *IDH* mutation status, we also assessed the degree to which our predicted molecular subtypes delineated survival trajectories, which is informative for gauging the potential utility of preoperative glioma classification models in clinical practice.

In summary, this is the first study to demonstrate that the integration of genetic risk profiles with MRI-based radiographic features may improve *IDH* status classification of glioma patients. Given the available data, our case study helps motivate future research on multimodal classifiers of clinically informative tumor features and highlights the potential to augment imaging-based ML algorithms for glioma with germline genetics.

## AUTHOR CONTRIBUTIONS

Conceptualization of project: TN and LK. Methodology: TN, GH, TB, RB, QZ, OG, and LK. Main analyses: TN, GH, and TB. Primary data collection and curation: TN, TB, SSF, GG, and LK. Drafting of the manuscript: TN, GH, and LK. All authors contributed to, reviewed and approved the final manuscript.

## ACKNOWLEDGEMENTS

This study received no funding.

## DATA AVAILABILITY

Genotype data of glioma cases from The Cancer Genome Atlas (TCGA) are available from the Database of Genotypes and Phenotypes (dbGaP) under accession phs000178. Radiomic data of glioma cases from TCGA can be obtained from The Cancer Imaging Archive (https://www.cancerimagingarchive.net)^24,25^. The data required for fitting polygenic risk scores for glioma are available at: https://zenodo.org/records/10790748.

