## Supplementary material for "Integration of MRI radiomics and germline genetics to predict the IDH mutation status of gliomas"

Affiliations:

**Supplementary Figure 1:** Overview of nested cross-validation procedure used for evaluation of IDH mutation classification in subset of 158 cases with genetic and imaging data.

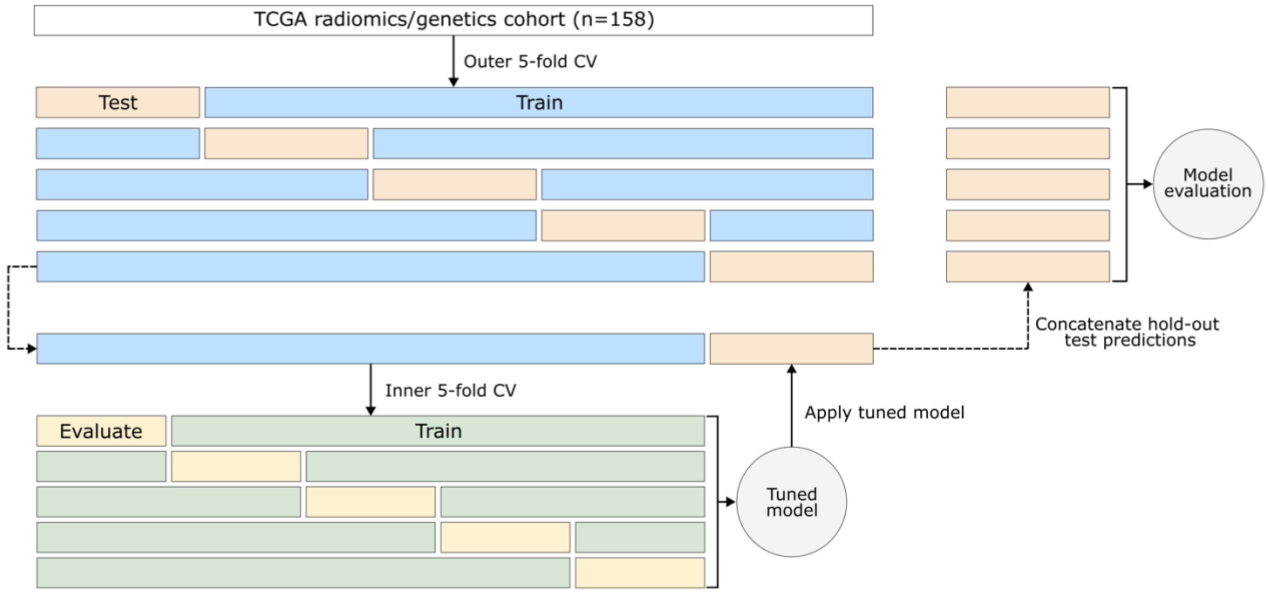

**Supplementary Figure 2: Comparison of distribution of AUC for classification of IDH mutation status across repeated nested cross-validations for single class models.** (**A**) Mean AUC and 95% confidence intervals (CI), approximated by the difference between the 97.5th and 2.5th percentiles of the AUC distribution across 500 iterations of the cross-validation procedure for each feature set. (**B**) Difference in mean AUC between model on y-axis and model on x-axis (i.e. $\Delta AUC=AUC_{y\text{-}axis}-AUC_{x\text{-}axis}$) for radiomic (Rad.), composite PRS (PRS), demographic (Dem.) and UNet-based autoencoder (UNET) features. Two-sided p-values testing the differences in AUC between each pair of models based on a resampled t-test are shown in parentheses.

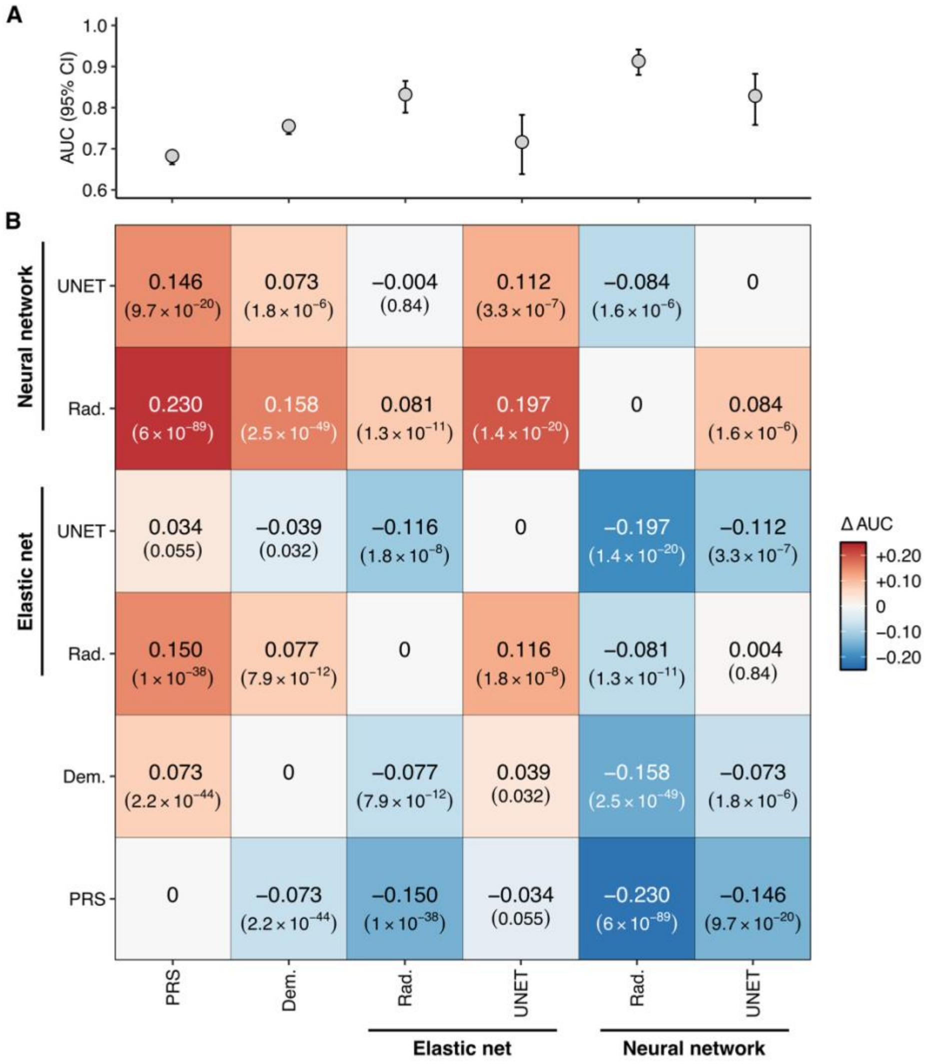

**Supplementary Figure 3: Comparison of distribution of AUC for classification of IDH mutation status across repeated nested cross-validations.** (**A**) Mean AUC and 95% confidence intervals (CI), approximated by the difference between the 97.5[th] and 2.5[th] percentiles of the AUC distribution across 500 iterations of the cross-validation procedure for each feature set. (**B**) Difference in mean AUC between model on y-axis and model on x-axis (i.e. $\Delta AUC = AUC_{y\text{-}axis} - AUC_{x\text{-}axis}$) for various combinations of radiomic (Rad.), composite PRS (PRS), demographic (Dem.) and UNet-based autoencoder (UNET) features. Two-sided p-values testing the differences in AUC between each pair of models based on a resampled t-test are shown in parentheses.

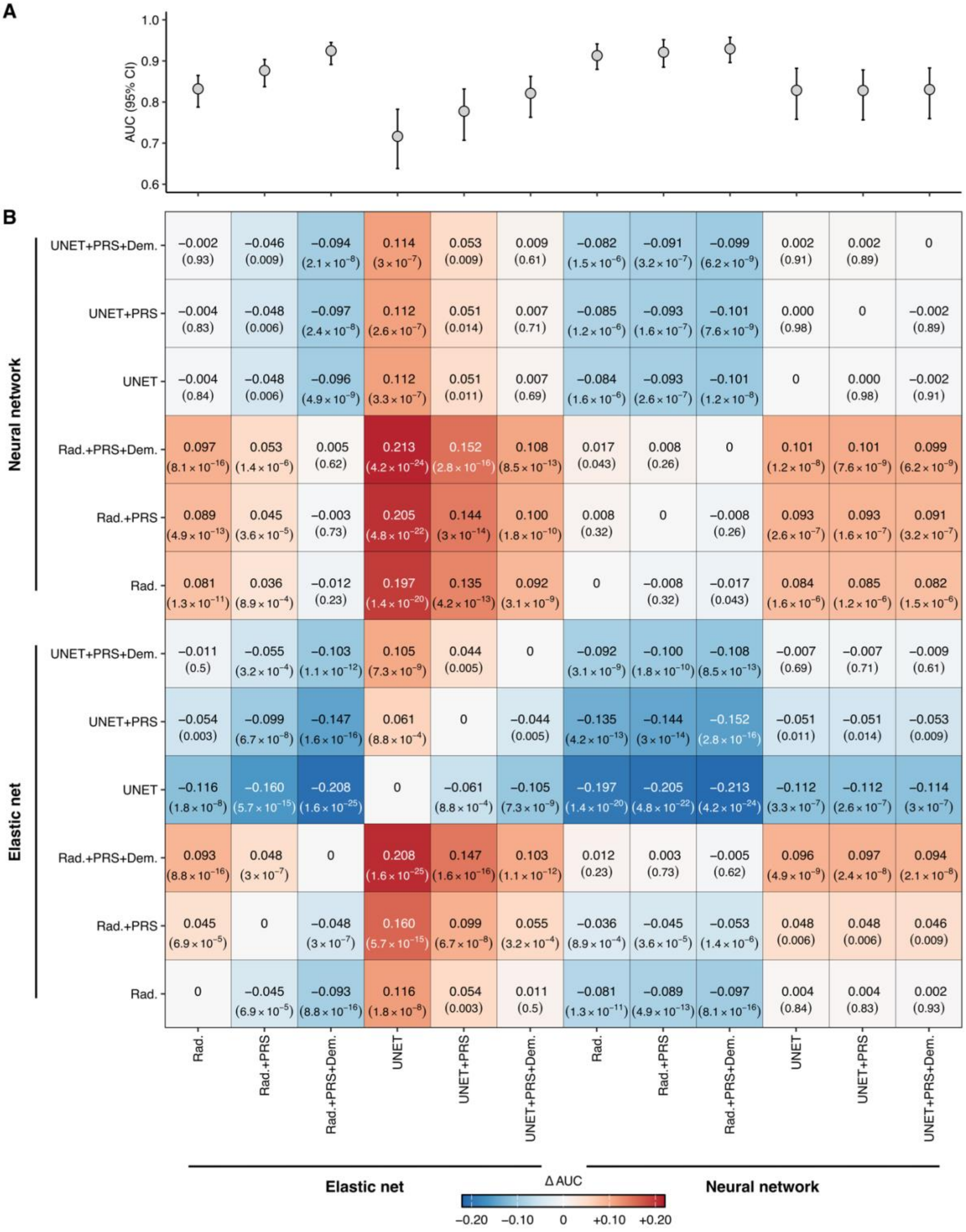

**Supplementary Figure 4: Distribution of AUC for classification of IDH mutation status from repeated nested cross-validation using radiomic features unadjusted for potential confounding factors.** AUC estimates from 500 random iterations of nested cross validation were obtained from IDH classification models using different classes of features. Demographics includes age at diagnosis and sex, whereas PRS refers to the composite PRS for IDH-mutant glioma.

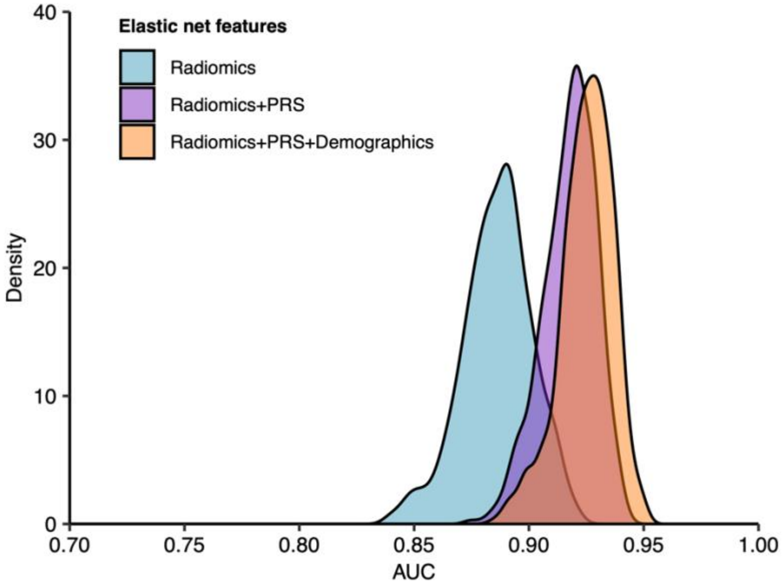

**Supplementary Table 1: Classification performance for IDH mutation status using radiomic features unadjusted for potential confounding factors.** Performance metrics are based on classification models for *IDH* mutation status in glioma cases. Models were trained with an elastic net. Demographics includes age at diagnosis and sex, whereas PRS refers to the composite PRS for IDH-mutant glioma. Two-sided p-values ($P_{\Delta AUC}$) testing the differences in AUC between the radiomics-based model and integrated models are based on a resampled t-test.

| Features | Accuracy | Precision | Recall | AUC (IQR) | | $P_{\Delta AUC}$ |
|---|---|---|---|---|---|---|
| Radiomics | 0.83 | 0.86 | 0.79 | 0.89 | (0.88, 0.90) | - |
| Radiomics + Demographics | 0.84 | 0.85 | 0.81 | 0.91 | (0.87, 0.93) | 0.032 |
| Radiomics + PRS | 0.86 | 0.88 | 0.81 | 0.92 | (0.89, 0.94) | $6.6\times10^{-5}$ |
| Radiomics + PRS + Demographics | 0.86 | 0.87 | 0.83 | 0.93 | (0.90, 0.94) | $1.2\times10^{-6}$ |

**Supplementary Table 2: Feature association with overall survival.** Features used for IDH status classification are examined for association with overall survival using univariate Cox proportional hazards regression.

| Feature | Feature category | HR (95% CI) | P value |
|---|---|---|---|
| Age at diagnosis | Demographic | 2.60 (1.93-3.51) | $4.40\times10^{-10}$ |
| Polygenic risk score (PRS) | Genetic | 0.84 (0.66-1.07) | 0.15 |
| NET T1Gd Bin 10 | Radiomic - Histogram | 1.34 (1.12-1.60) | 0.0016 |
| NET FLAIR Bin 4 | Radiomic - Histogram | 1.06 (0.85-1.33) | 0.61 |
| NET T1Gd Bin 1 | Radiomic - Histogram | 1.02 (0.81-1.28) | 0.87 |
| NET T2 Bin 10 | Radiomic - Histogram | 1.01 (0.81-1.26) | 0.92 |
| NET T1 Bin 7 | Radiomic - Histogram | 0.96 (0.77-1.20) | 0.73 |
| NET T1Gd Bin 6 | Radiomic - Histogram | 0.95 (0.74-1.21) | 0.67 |
| NET T1Gd Bin 3 | Radiomic - Histogram | 0.91 (0.72-1.15) | 0.43 |
| NET FLAIR Bin 9 | Radiomic - Histogram | 0.87 (0.67-1.14) | 0.32 |
| NET T1Gd Bin 5 | Radiomic - Histogram | 0.75 (0.57-0.99) | 0.046 |
| Mean NET T1Gd | Radiomic - Intensity | 1.42 (1.12-1.80) | 0.0043 |
| STD NET T2 | Radiomic - Intensity | 1.33 (1.03-1.71) | 0.029 |
| Mean NET T1 | Radiomic - Intensity | 1.32 (1.04-1.68) | 0.02 |
| Parietal | Radiomic - Spatial | 1.12 (0.91-1.38) | 0.29 |
| Frontal | Radiomic - Spatial | 0.73 (0.57-0.94) | 0.016 |
| GLOBAL NET FLAIR Kurtosis | Radiomic - Texture | 1.32 (1.09-1.60) | 0.0045 |
| GLCM NET T1 Correlation | Radiomic - Texture | 1.28 (0.92-1.78) | 0.15 |
| GLRLM NET T2 SRE | Radiomic - Texture | 1.20 (0.95-1.51) | 0.13 |
| GLOBAL NET FLAIR Skewness | Radiomic - Texture | 0.99 (0.76-1.30) | 0.97 |
| GLCM NET T2 Homogeneity | Radiomic - Texture | 0.94 (0.75-1.16) | 0.55 |
| GLRLM NET T2 LRLGE | Radiomic - Texture | 0.80 (0.60-1.05) | 0.1 |
| GLOBAL NET FLAIR Variance | Radiomic - Texture | 0.77 (0.64-0.93) | 0.0067 |
| NGTDM NET T1Gd Busyness | Radiomic - Texture | 0.75 (0.55-1.02) | 0.069 |
| GLCM NET T1Gd AutoCorrelation | Radiomic - Texture | 0.75 (0.59-0.94) | 0.014 |
| GLCM NET T1Gd SumAverage | Radiomic - Texture | 0.74 (0.59-0.93) | 0.0089 |
| NGTDM NET FLAIR Busyness | Radiomic - Texture | 0.73 (0.56-0.97) | 0.028 |
| GLCM NET FLAIR Variance | Radiomic - Texture | 0.73 (0.58-0.91) | 0.0052 |
| NGTDM NET T1 Contrast | Radiomic - Texture | 0.70 (0.52-0.93) | 0.013 |
| GLRLM NET FLAIR HGRE | Radiomic - Texture | 0.69 (0.56-0.84) | $2.70\times10^{-4}$ |
| NGTDM NET T1 Busyness | Radiomic - Texture | 0.56 (0.40-0.81) | 0.0016 |
| GLSZM NET T1 LZLGE | Radiomic - Texture | 0.43 (0.26-0.72) | 0.0011 |
| GLSZM NET FLAIR LZLGE | Radiomic - Texture | 0.29 (0.13-0.69) | 0.0051 |
| ET/TC | Radiomic - Volumetric | 1.51 (1.22-1.87) | $1.30\times10^{-4}$ |
| ET/WT | Radiomic - Volumetric | 1.30 (1.08-1.57) | 0.005 |
| NET/BRAIN | Radiomic - Volumetric | 0.70 (0.52-0.94) | 0.016 |
| NET/TC | Radiomic - Volumetric | 0.66 (0.54-0.82) | $1.30\times10^{-4}$ |
| NET/WT | Radiomic - Volumetric | 0.64 (0.49-0.83) | $8.40\times10^{-4}$ |
| SOLIDITY NET | Radiomic - Volumetric | 0.61 (0.49-0.77) | $3.30\times10^{-5}$ |
| Center of gravity (y-axis) | Radiomic - Tumor Growth Model | 1.31 (1.03-1.66) | 0.028 |

Abbreviations: NET=Non-enhancing tumor, WT=Whole tumor, TC=Tumor core, ET=Enhancing tumor, GLCM= Grey-level co-occurrence matrix, GLSZM=Gray-level size zone matrix, GLRLM= Gray-level run-length matrix, SZLGE=Small zone low gray-level emphasis, SRE=Short run emphasis, NGTDM=Neighborhood grey-tone difference matrix
